# A robust framework for harmonising health measures across international cohorts: Evidence from the COVID-19 pandemic

**DOI:** 10.1101/2025.09.09.25335409

**Authors:** James Lian, Pedro F Zuccolo, Omid V. Ebrahimi, Daniel Fatori, Abhaya Adlakha, Juan F. De La Hoz, Younga H. Lee, Adriana Carneiro, Isabela M. Benseñor, Paulo A. Lotufo, Alessandra C. Goulart, Justin D. Tubbs, Devon Watts, Yu Zhou, Lorenza Dall’Aglio, Mihael Cudic, Morgane Kuenzi, CGMHC Consortium, Ronald C. Kessler, Vikram Patel, André Brunoni, Jordan W. Smoller, Sarah Bauermeister

## Abstract

Cross-national research on health trajectories requires harmonised measures that are valid and comparable. However, measurement scales often differ between cohorts in item content and cultural context. We present a structured framework to harmonise mental health measures, using depression and anxiety symptoms across two longitudinal cohorts that used different self-report measures with heterogeneous item codings: ELSA-UK and ELSA-Brasil.

Data were collected before, during, and after the COVID-19 pandemic, providing a natural experiment for examining temporal changes in mental health. We applied a theory-driven strategy to align item content across cohorts relying on a priori assumptions of cross-national item equivalence and binary comparability. This involved: (1) mapping items to DSM-5 symptom domains via expert review; (2) transforming response formats into harmonised ordinal indicators; (3) leveraging the Harmony AI tool to identify semantically equivalent items; and (4) establishing measurement invariance across waves and cohorts using multi-group confirmatory factor analysis (MGCFA).

Within this framework, scalar invariance was achieved for depression in both longitudinal and cross-cohort models, enabling meaningful latent mean comparisons between cohorts. For anxiety, scalar invariance was supported within but not across cohorts, likely due to the limited number of conceptually matching items. These findings highlight that the success of harmonisation relies on the quality and conceptual alignment of available items. Our results demonstrate the feasibility of robust cross-cultural comparisons and provide a methodological template for future harmonisation efforts in global health research.

## Introduction

Internationally harmonised health data have become increasingly essential for addressing key global health research questions [1]. In epidemiological research, meaningful comparisons across countries, time points, and populations rely on the availability of harmonised measures [2,3]. Harmonisation enables pooled analyses, enhances statistical power, and facilitates generalisability of findings. It also expands the range of contextual and demographic variation in exposures, such as socioeconomic status or public health policies, by integrating diverse samples from multiple settings [4–6]. However, in the field of mental health, where assessment tools often vary in content, structure, and cultural interpretation, creating comparable measures across studies remains a significant methodological challenge [7].

Several initiatives have published guidelines to promote standardised harmonisation practices [4] and successful examples are emerging [2,4,8–10]. Despite this progress, harmonising mental health data across heterogeneous cohorts remains challenging. In many cases, harmonisation is restricted to studies using identical instruments, such as the HRS International Family of Studies, which employs the CES-D across member cohorts [11]. When cohorts rely on different instruments, harmonisation strategies typically include retrospective item mapping, variable recoding, or the application of item response theory (IRT) [12,13]. These strategies, while valuable, face limitations: they may depend heavily on subjective decisions about item equivalence, impose strong assumptions such as unidimensionality, or fail to evaluate whether constructs are measured equivalently across time and groups (3,14).

To address these limitations, researchers have increasingly called for harmonisation frameworks that explicitly test measurement equivalence across diverse populations (Ebrahimi et al., 2024). Multi-group confirmatory factor analysis (MGCFA) offers such a framework, providing a rigorous psychometric method for assessing whether latent constructs are measured consistently across cohorts, cultures, and timepoints [15]. MGCFA involves fitting a series of increasingly constrained models — configural, metric, and scalar invariance — to evaluate whether observed differences reflect genuine variation in underlying constructs rather than artefacts of measurement bias or cultural interpretation. Importantly, MGCFA offers a scalable and statistically principled foundation for global harmonisation of health constructs, particularly when studies differ in language, format, or item selection (Putnick & Bornstein, 2016; Van de Schoot et al., 2015).

Although prior work has examined measurement invariance either longitudinally or cross-sectionally, few harmonisation strategies address both dimensions simultaneously (Ebrahimi, 2024). Without evidence that a measure functions consistently across time and across populations, observed differences may conflate true change with artefacts of measurement [16]. Moreover, standardisation of scales between cohorts with different instruments is rarely attempted in a way that also explicitly assesses the similarity of item functioning. To our knowledge, no prior study has combined expert-driven item mapping, data-driven semantic matching, harmonisation of heterogeneous response formats, and formal invariance testing across both time and cohorts in a single methodological pipeline.

The COVID-19 pandemic provides a unique natural experiment for testing harmonisation methods due to its global reach, profound psychological impact, and variability in public health responses. While extensive research has documented health effects during the pandemic [17–22], relatively little is known about cross-national differences in mental health trajectories during this period. Countries varied widely in the timing, duration, and stringency of containment policies [23], contributing to variability in mental health responses. Vulnerable groups, such as women, young people, and individuals from socioeconomically disadvantaged backgrounds, were disproportionately affected [24–29]. Moreover, mental health trends fluctuated over time, often peaking during initial lockdowns and attenuating thereafter [30–36]. These unique circumstances provide an opportunity to test whether harmonisation methods can yield valid, comparable mental health indicators across countries with different instruments, contexts, and pandemic responses.

## Objectives

As part of the COVID Global Mental Health Consortium (CGMHC), we aimed to develop and apply a systematic harmonisation framework to generate depression and anxiety measures that are comparable both longitudinally and cross-culturally, despite differences in item content and response formats across instruments. Data came from two culturally and socioeconomically distinct cohorts: the English Longitudinal Study of Ageing (ELSA-UK) [37] and the Brazilian Longitudinal Study of Ageing (ELSA-Brasil) [38,39]. Using these studies, we implemented a multi-step process combining expert item mapping, AI-based semantic similarity matching, and multi-group confirmatory factor analysis (MGCFA) to evaluate measurement invariance across time and cohorts. Our objective was to test the feasibility of this framework by deriving harmonised latent factor scores for depression and anxiety, and to highlight both its potential and its limitations for future cross-national health research.

## Methods

### Data sources

#### Participating cohorts

This study draws on two longitudinal population-based cohorts: the ELSA-Brasil COVID-19 Mental Health Cohort study, and the English Longitudinal Study of Ageing (ELSA-UK). Despite their similar names, these studies differ substantially in measurement instruments, cultural context, and data collection procedures.

The ELSA-Brasil cohort is a prospective study of 15,105 participants from six major Brazilian cities, established to investigate the clinical and sociodemographic determinants of chronic diseases and mortality in a middle-income country context [38]. Participants are university employees (active or retired), aged 35-74 years, and free from major neurocognitive disorders at enrolment. Nested within the larger study is ELSA Brasil COVID-19. During the pandemic, participants from the São Paulo site (n = 4,191) were invited to complete online assessments of mental health, performed in four waves: c1 (May–July 2020), c2 (July– September 2020), c3 (October–December 2020), and c4 (April–June 2021) [28,39]. A total of 2,691 individuals who completed at least one wave were included in the analysis.

The ELSA-UK cohort, launched in 2002, follows a nationally representative sample of adults aged 50 and older living in England, tracking health, social, and economic changes in later life. For this study, we used data from wave 9 (2018–2019), two COVID-19 sub-studies (June– July 2020 and November–December 2020), and wave 10 (2021–2023), comprising 11,004 participants. For both cohorts, we selected waves from 2018–2023 that were available in the DPUK portal and contained comparable mental health measures, rather than aligning assessments by calendar date. This ensured that harmonisation focused on measurement equivalence, even though data collection periods differed between countries. Figure 1 depicts the timing of assessments included in these analyses. Ethical approval was obtained from relevant local committees for all waves and cohorts.

**Figure 1.**
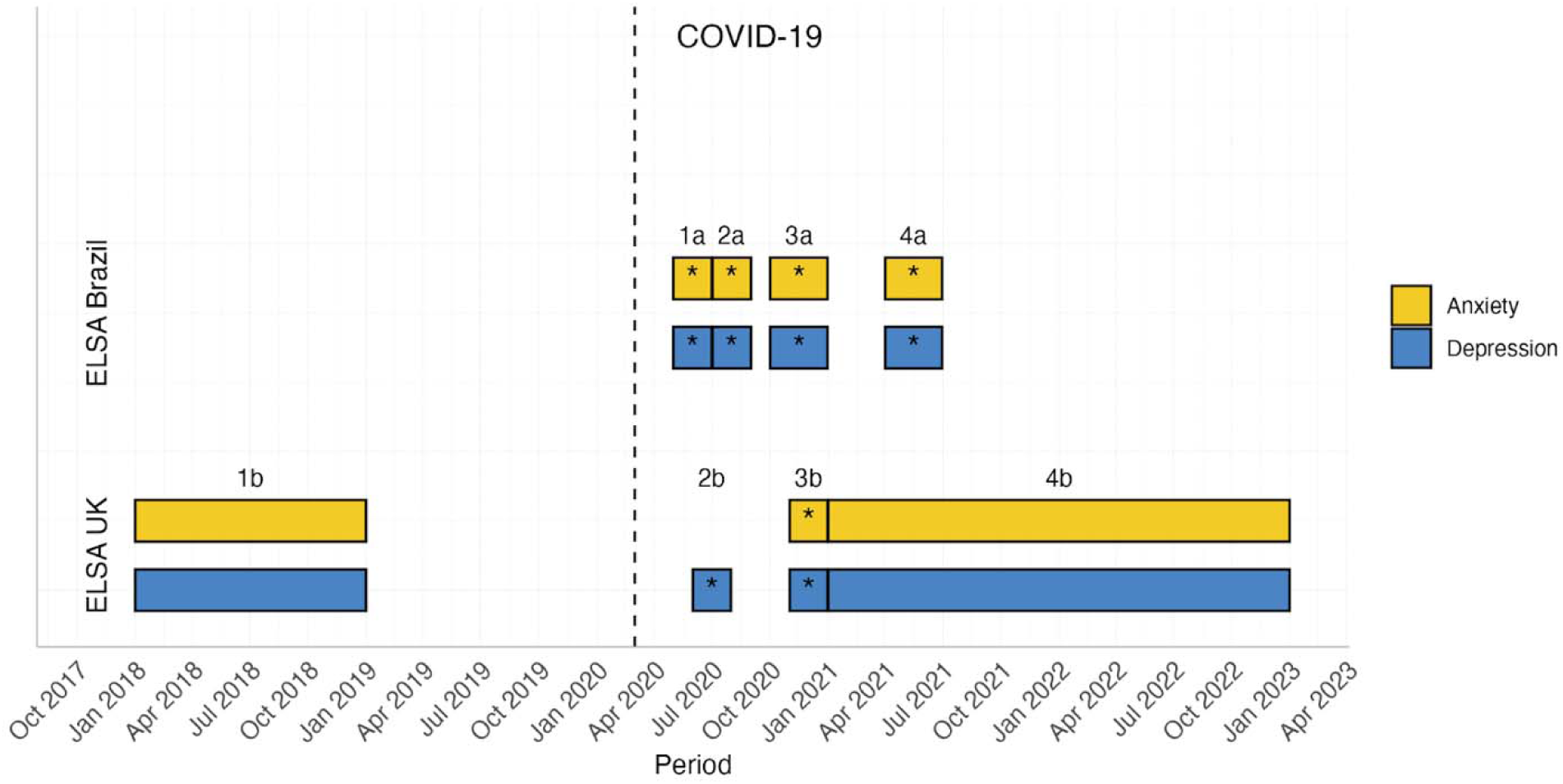
Timing of data collection around the COVID-19 pandemic. *Note.* ELSA Brasil waves (N = 2691): **1a)** May–July 2020**, 2a)** July–September 2020, **3a)** October–December 2020, and **4a)** April to June 2021. ELSA UK waves (N = 11,004): **1b)** 2018-2019, **2b)** June-July 2020, **3b)** November, December 2020, and **4b)** 2021-2023.

#### Setting

All data were accessed and analysed via the Dementias Platform UK (DPUK) Data Portal, a secure, cloud-based research environment equipped with pre-installed statistical tools [40]. DPUK can only be accessed by approved users and data is never downloaded or analysed outside this environment. R version 4.1 was used for analysis. Analyses were conducted between December 2024 and May 2025.

#### Mental health measures

Mental health symptoms in the ELSA-Brasil COVID-19 study were assessed using the Brazilian version of the Depression, Anxiety, and Stress Scale – 21 items (DASS-21) [41]. This instrument was designed to measure symptoms related to depression, anxiety, and stress in the past week. It consists of 21 self-report items rated on a four-point Likert scale (0 = “strongly disagree” to 3 = “totally agree”), based on the frequency or intensity of experienced symptoms. The scale includes three subscales—depression, anxiety, and stress—each composed of seven items; scores can be computed separately for each domain or summed into a total score (range: 0–63), with higher values indicating greater symptom severity. Questions from DASS-21 were asked in Brazilian Portuguese and were translated to English for this study.

Mental health in the ELSA UK cohort was assessed using the 8-item version of the Center for Epidemiologic Studies Depression Scale (CES-D-8) [42], the Generalized Anxiety Disorder Scale (GAD-7) [43], the Control, Autonomy, Self-Realization and Pleasure Scale (CASP-12) [44], and a four-item personal well-being questionnaire (ONS-4) [45].

The CES-D-8 is a brief self-report scale designed to measure the frequency of depressive symptoms over the past week. In contrast to the original CES-D-20 that is rated on a four-point Likert scale, the CES-D-8 uses a dichotomous (yes/no) format to reduce participant burden and confusion [46]. The GAD-7 aims to evaluate symptoms of generalised anxiety disorder over the past two weeks. It includes 7 items rated on a four-point Likert scale (0 = “not at all” to 3 = “nearly every day”). The total score ranges from 0 to 21, with higher scores reflecting more severe anxiety symptoms. The CASP-12 was developed to assess quality of life in older adults, focusing on control, autonomy, self-realisation, and pleasure. It comprises 12 items rated on a four-point Likert scale (1-4, Often-Never, with some items negatively worded and reversed when computing scores), where higher scores indicate better quality of life. The ONS-4 assesses personal well-being using four measures: Life satisfaction, Worthwhile (i.e., subjective sense of purpose and meaning in life), Happiness, and Anxiety. The four questions are on an 11-point scale from 0 to 10, where 0 is “not at all” and 10 is “completely”.

### Harmonisation pipeline

To facilitate cross-cohort comparisons, a team of subject matter experts (PFZ, JL, DF, and AA) conducted a 5-step process of retrospective harmonisation of mental health measures, as shown in Figure 2. The experts are either clinical psychologists and/or hold doctorates in psychiatric epidemiology.

**Figure 2.**
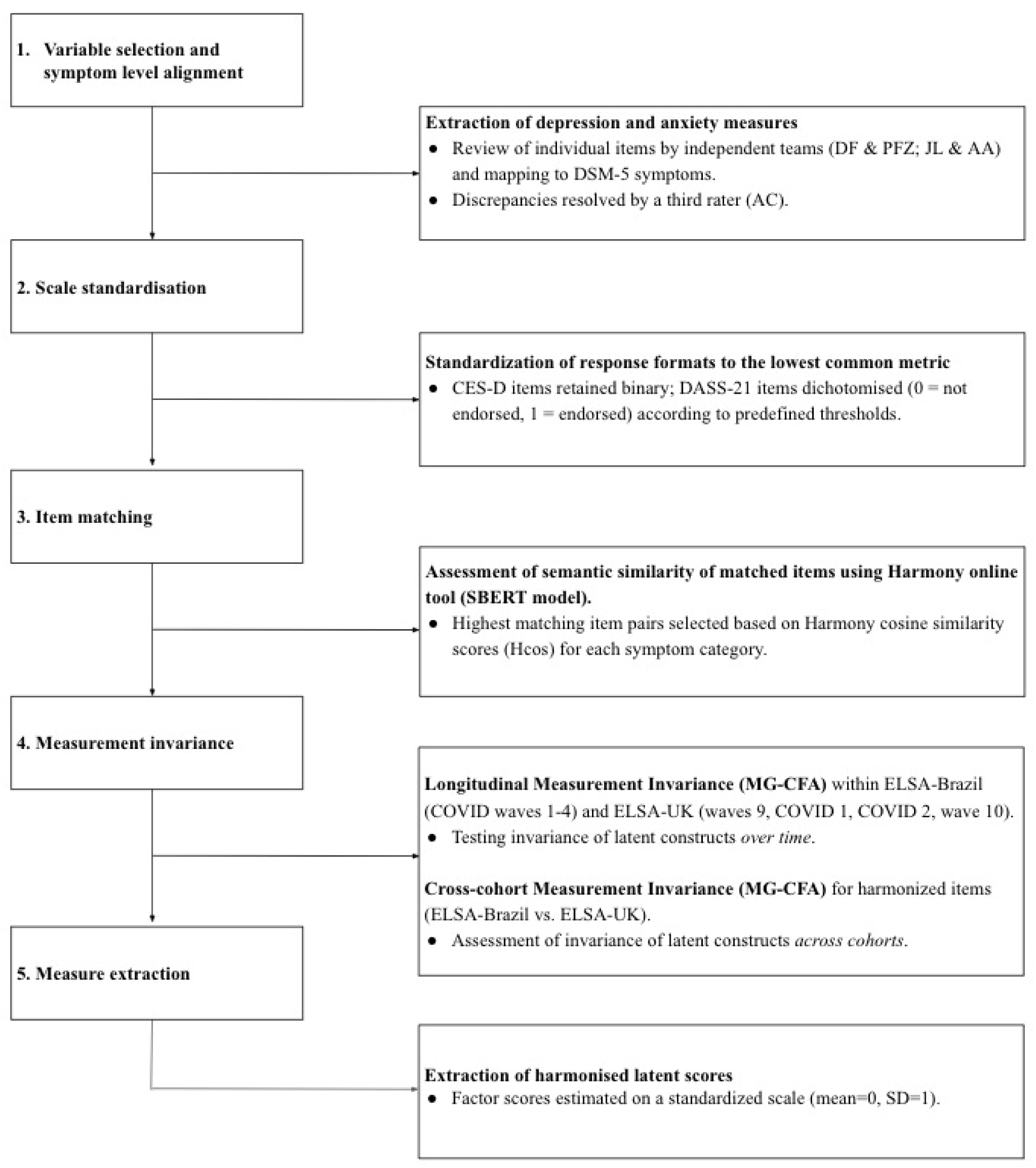
Harmonisation pipeline.

#### 1. Variable selection and symptom level alignment

Data dictionaries from each cohort were reviewed to identify and extract all measures assessing depression and anxiety. Symptom categories for these conditions were defined based on DSM-5-TR criteria for major depressive disorder and generalised anxiety disorder. All relevant items were then compiled in a spreadsheet to allow for symptom-level and item-level alignment (see supplementary).

Two independent teams (DF and PFZ; JL and AA) reviewed the individual items of each scale and mapped them to specific depression and anxiety symptom categories (e.g., “fatigue”, “low energy”; or “worry”, or “irritability”). Items were considered similar if they used comparable wording (e.g., “I felt that life was meaningless” and “I feel that my life has meaning”) or if they were interpreted as referring to the same symptom (e.g., “I felt down-hearted and blue” and “I felt depressed”). Items that were judged to capture multiple symptoms (e.g., “depressed mood” and “anhedonia”) were coded under all applicable categories.

Discrepancies between the two groups were resolved by a third independent rater with extensive clinical and psychometric experience (AC) (see supplementary).

#### 2. Scale standardisation

A major challenge in the harmonisation process was the variation in the number of response levels across instruments (binary or likert-scale). For example, the CES-D (used in ELSA-UK) adopted a binary response format (e.g., 0 = “No”, 1 = “Much of the time during the past week”), while the DASS-21 (used in ELSA-Brasil) relied on a four-point Likert scale (from 0 = “Did not apply at all” to 3 = “Applied a lot, or most of the time”). To enable cross-cohort comparability, we adopted a conservative approach by aligning all items to the lowest common response structure, which led to all items being binarised. Specifically, CES-D items were retained as binary format (1 = “Much of the time”), whereas DASS-21 items were dichotomised using a predefined threshold (Table 1).

**Table 1.** Item recoding thresholds for DASS, CES-D, CASP, GAD, and QOL questionnaires.

| DASS |  | CES-D |  | CASP |  | GAD |  | QOL |  |
| --- | --- | --- | --- | --- | --- | --- | --- | --- | --- |
| Value | Label | Value | Label | Value | Label | Value | Label | Value | Label |
| 0 | Did not apply at all | 0 | Not much of the time during the past week | 1 | - | 0 | Never | 0 | - |
| 1 | Applied to some degree, or for a short time | 1 | Much of the time during the past week | 2 | - | 1 | Several days | 1 | - |
| 2 | Applied to a considerable degree, or for a large part of the time |  |  | 3 | - | 2 | More than half of the days | 2 | - |
| 3 | Applied a lot, or most of the time |  |  | 4 | - | 3 | Almost every day | 3 | - |
|  |  |  |  |  |  |  |  | 4 | - |
|  |  |  |  |  |  |  |  | 5 | - |
|  |  |  |  |  |  |  |  | 6 | - |
|  |  |  |  |  |  |  |  | 7 | - |
|  |  |  |  |  |  |  |  | 8 | - |
|  |  |  |  |  |  |  |  | 9 | - |
|  |  |  |  |  |  |  |  | 10 | - |

*Note.* Dotted line represents the cut-off for recoding, where items above the dotted line were recoded to 0, and items below the dotted line are recoded to 1. A dash represents a numeric scoring system where labels aren't used.

#### 3. Item matching

To fit MGCFA models, each cohort must have an equal number of items representing each construct (e.g., depression and anxiety), although the specific item wording can differ if the items are conceptually comparable. We used the Harmony online harmonisation tool [2,47] to assess semantic similarity between items across ELSA-UK and ELSA-Brasil. Harmony uses the Sentence Bidirectional Encoder Representations from Transformers model (SBERT) model [48], a deep learning model optimised for natural language inference, to calculate cosine similarity which measures the cosine of the angle between their embedding vectors in multidimensional space, with values closer to 1 indicating that the items are more semantically alike, and values closer to 0 indicating little semantic overlap.

We did not apply a rigid minimum Hcos threshold for inclusion because high semantic similarity scores do not always guarantee conceptual equivalence in a clinical/psychometric context, and vice versa. Instead, the Harmony scores were used as a starting point to identify candidate matches, which were then reviewed by domain experts against DSM-5 symptom definitions and the broader measurement context (e.g., scale format, intended construct). This review allowed the inclusion of some lower-scoring pairs (e.g., Hcos ≈ 0.30 for “lack of motivation or apathy”), when the items were judged to represent the same symptom domain despite differences in wording or translation nuance. Conversely, some higher-scoring pairs were excluded if they were judged to differ conceptually (e.g., “not feeling much worth as a person” vs. “not feeling one’s life is worthwhile”).

Where multiple candidate items were available for the same symptom category, the highest-scoring pair that also met expert conceptual criteria was selected to construct the harmonised scale for each cohort.

#### 4. Measurement invariance testing

After selecting harmonised items, we tested whether the depression and anxiety constructs were measured equivalently across cohorts and over time using multi-group confirmatory factor analysis (MGCFA). Measurement invariance evaluates whether the relationships between latent constructs (e.g., depression) and their observed indicators are comparable (i) longitudinally within each cohort and (ii) cross-sectionally between cohorts. Without establishing invariance, any observed differences between groups may reflect measurement artefacts rather than true differences in the latent construct [14,49].

MGCFA proceeds by fitting a sequence of increasingly constrained models:

1. *Configural invariance* tests whether groups share the same underlying factor structure, that is, whether the same number of factors and the same set of items load on each factor. This serves as a baseline for further invariance testing.
2. *Metric invariance* constrains the unstandardised factor loadings (metric coefficients) to be equal across groups. This ensures that a one-unit change in the latent variable corresponds to the same expected change in each observed item in all groups, establishing a common measurement scale. Standardised loadings may still differ because of differences in item or factor variances [50,51]. Achieving metric invariance permits valid comparisons of associations involving the latent construct (e.g., regressions).
3. *Scalar invariance* adds equality constraints on item thresholds (for categorical indicators) or intercepts (for continuous indicators), ensuring that group differences in observed means reflect true differences in the latent construct rather than measurement bias. Scalar invariance is required to compare latent means [52].
4. *Strict invariance* further constrains residual variances to be equal across groups. While difficult to achieve, it is useful in certain contexts, such as when comparing factor means from dichotomous items [53,54].

When full invariance cannot be established, *partial measurement invariance (PMI)* can be applied by relaxing specific constraints (e.g., freeing thresholds for some items) while retaining others, provided that at least two invariant items anchor each factor [49]. The highest level of invariance attained dictates the types of valid cross-group comparisons: configural and metric invariance allow for comparing associations, whereas scalar (full or partial) invariance is required for comparing latent means (Yoon & Kim, 2014).

#### 5. Harmonised measure extraction

Once partial or full measurement invariance was established, we applied a two-step procedure to derive harmonised latent scores for subsequent analyses. These scores served as cross-cohort, longitudinally comparable measures of depression and anxiety.

Factor scores estimate an individual’s standing on an underlying latent construct, typically on a standardised scale (mean = 0, SD = 1). When measurement invariance holds, these scores can be meaningfully compared across groups and time points. Scores at the distributional extremes indicate stronger associations with the latent factor, whereas values near zero reflect weaker symptom endorsement. By constraining measurement parameters across groups and waves during estimation, scores are placed on a shared metric, thereby supporting valid comparisons of mental health trajectories across methodologically distinct cohorts [55]

### Statistical analyses

First, we assessed longitudinal measurement invariance within each cohort separately: ELSA-Brasil (COVID waves 1–4) and ELSA-UK (waves 9, COVID 1, COVID 2, and wave 10). For each construct, we tested a sequence of increasingly constrained models: (1) a configural model to examine whether the same factor structure held across waves; (2) a metric model to assess equality of unstandardised factor loadings; and (3) a scalar model to evaluate equality of both loadings and item thresholds across time.

We then tested cross-cohort measurement invariance using MGCFA on the wave-specific item sets retained from the longitudinal models. This ensured that the cross-cohort models were informed by the longitudinal invariance results. For each construct, configural, metric, and scalar models were fitted across the two cohorts using identical matched item sets. Cross-cohort models were estimated for each wave separately rather than collapsing or averaging across time points, allowing us to examine invariance within specific temporal contexts.

Model fit was evaluated using standard CFA fit indices: RMSEA, CFI, TLI, and SRMR [56]. Following Hu and Bentler’s two-index approach, acceptable fit was defined as RMSEA ≤

.06; SRMR ≤ .08; CFI/TLI ≥ .95 with values of CFI/TLI > .90 considered adequate [57]. All models were estimated in *lavaan* (Rosseel, 2012) using the WLSMV estimator, which is suitable for binary indicators [58]. Analysis scripts are available at: https://github.com/cgmhc/cgmhc_harmonisation.

## Results

Table 2 presents baseline descriptive statistics for each cohort. The mean age was 69 years in ELSA-UK and 61 years in ELSA-Brasil. Educational attainment was higher in ELSA-Brasil, with 95% of participants educated at high school level or above, compared with 70% in ELSA-UK.

**Table 2.** Descriptive statistics of demographics across ELSA-UK and ELSA-Brasil cohorts.

| Cohort | ELSA-UK (N=7361) | ELSA-Brasil (N=2,636) |
| --- | --- | --- |
| Baseline year | 2020 | 2020 |
| Age (years, SD) | 69 (10) | 61.3 (8.5) |
| Female sex, n (%) | 4152 (56%) | 1507 (57.2%) |
| Education, n (%) | Below High School = 984 (13%)<br>High school or higher = 5110 (70%)<br>Missing = 1267 (17%) | Below High School = 68 (2.6%)<br>High School or higher = 2498 (94.8%)<br>Missing = 70 (2.6%) |
| Ethnicity (Ethnic minority), n (%) | 348 (5%) | 853 (32.8%) |
| Employment (Retired), n (%) | 4660 (63%) | 489 (24.1%) |
| Household (Living alone), n (%) | 1620 (22%) | 329 (16.8%) |

Table 3 summarises the Harmony item-matching process. Five matching item pairs were identified for depression (depressed mood, anhedonia, worthlessness, hopelessness, apathy) and four for anxiety (worry, general anxiety, irritability, restlessness). Cosine similarity (Hcos) scores varied across matches, with final selections based on both semantic similarity and conceptual alignment with DSM-5 symptom domains. Cut-offs used to standardise and recode response categories across cohorts are shown in Table 1.

**Table 3.**
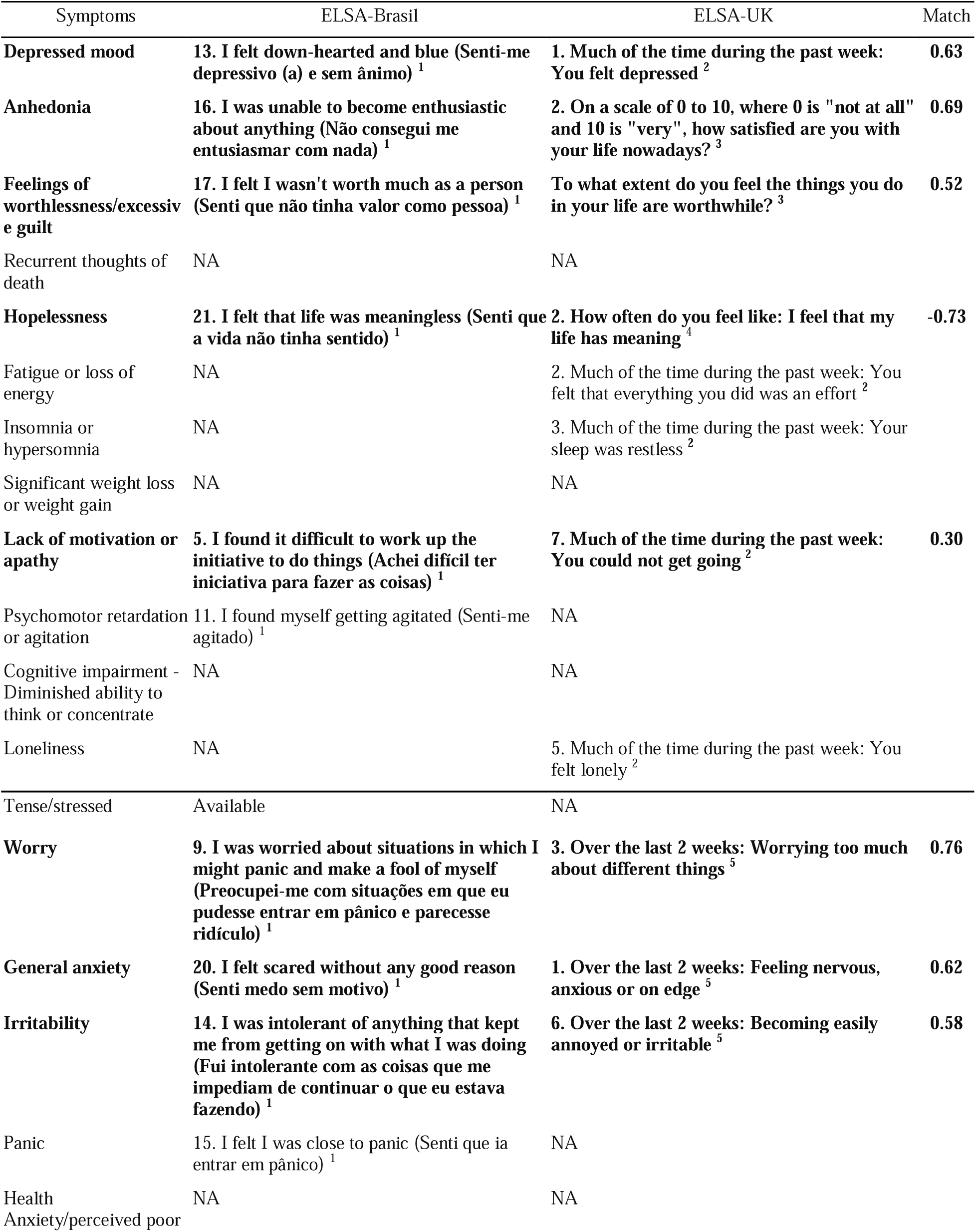
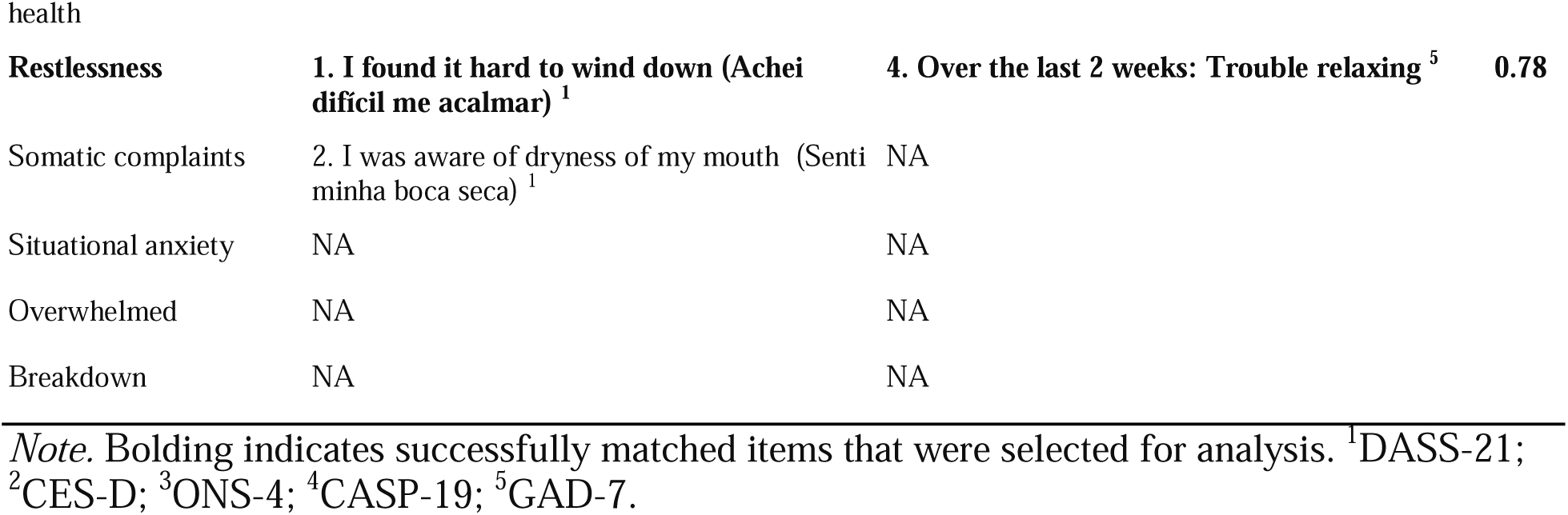
Items matched across cohorts by semantic similarity using Harmony tool.

Table 4 shows longitudinal measurement invariance results within each cohort. In ELSA-Brasil, configural, metric, and scalar models for both depression and anxiety demonstrated excellent fit (RMSEA < .06, CFI/TLI > .95), supporting full scalar invariance across four COVID-19 waves. In ELSA-UK, all depression models showed acceptable fit, with only a small decline in fit indices from configural to scalar models. Anxiety models demonstrated excellent fit at all levels of invariance, supporting full scalar invariance across time.

**Table 4.**
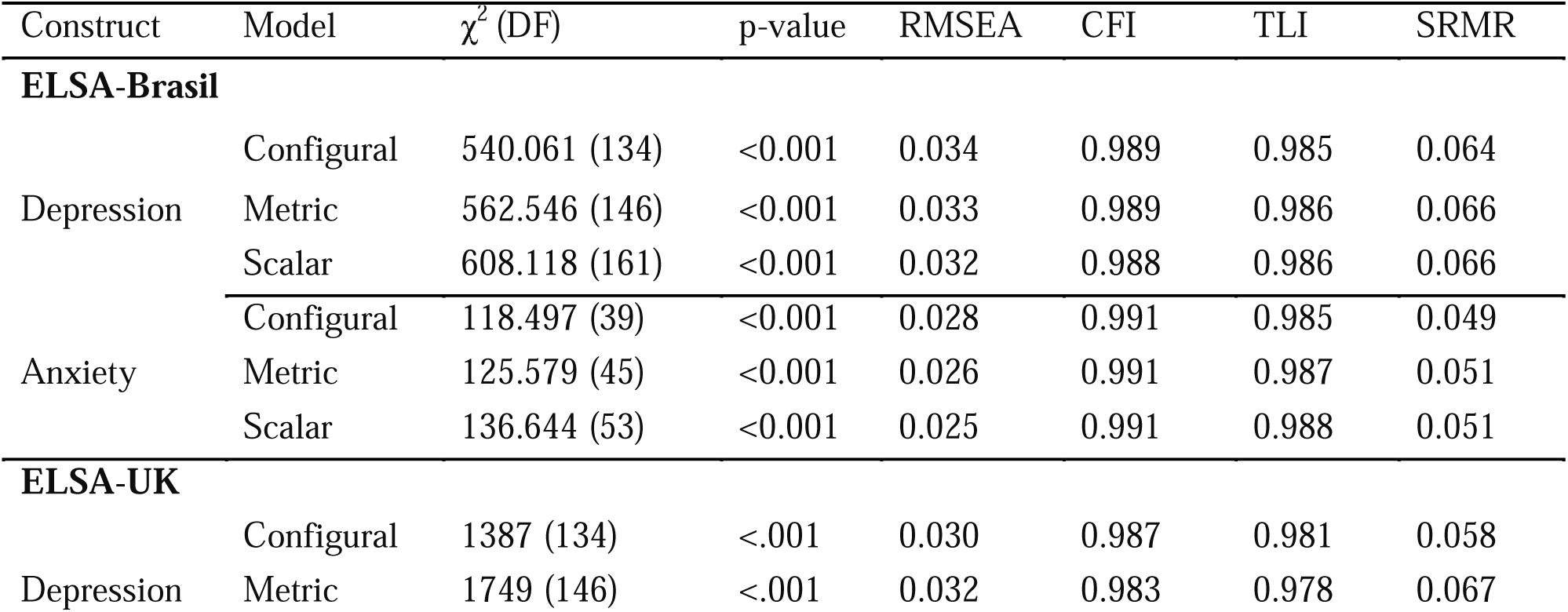
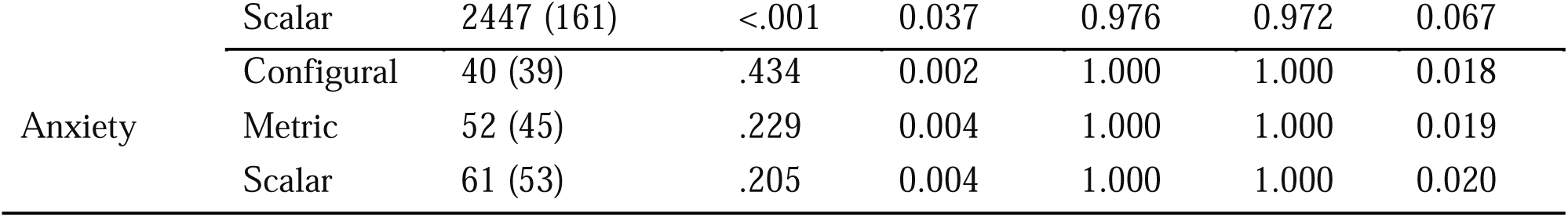
Fit indices for longitudinal measurement invariance of depression and anxiety within ELSA-Brasil (N = 2,636) and ELSA-UK (N = 13,695) cohorts.

Table 5 summarises cross-cohort invariance testing. For depression, acceptable fit was maintained from configural through scalar models, indicating that latent mean comparisons between ELSA-UK and ELSA-Brasil are valid. For anxiety, configural and metric models fit well, but the scalar model failed to converge, suggesting that latent mean comparisons for anxiety across cohorts may be biased by measurement non-equivalence.

**Table 5.** Fit indices for cross-cohort measurement invariance models of depression and anxiety in ELSA-Brasil (n=2,636) and ELSA UK cohorts (N = 13,695).

| Construct | Model | $\chi^2$ (DF) | p-value | RMSEA | CFI | TLI | SRMR |
| --- | --- | --- | --- | --- | --- | --- | --- |
| **Depression** | Configural | 1926.638 (268) | <.001 | .031 | .988 | .982 | .060 |
|  | Metric | 2676.207 (296) | <.001 | .035 | .982 | .977 | .070 |
|  | Scalar | 3219.805 (307) | <.001 | .038 | .978 | .973 | .068 |
| **Anxiety** | Configural | 158.297 (78) | <.001 | .013 | .999 | .998 | .025 |
|  | Metric | 212.696 (93) | <.001 | .015 | .999 | .998 | .028 |
|  | Scalar | - | - | - | - | - | - |

## Discussion

Our study illustrates the feasibility of retrospectively harmonising psychological symptom measures across culturally distinct ageing cohorts using a structured, theory-driven approach. We combined expert item mapping with AI-enabled semantic matching and MGCFA with the aim to harmonise cohorts with substantial differences in item content, response scales, and cultural context. In summary, we were able to derive harmonised depression and anxiety constructs from the ELSA-UK and ELSA-Brasil longitudinal studies. Crucially, we found that valid latent depression scores could be estimated comparably across time and between countries. Within each cohort, both depression and anxiety showed full longitudinal measurement invariance, indicating that the underlying constructs were measured equivalently over successive waves. Moreover, our cross-cohort analysis demonstrated scalar invariance for depression across the UK and Brazil samples, implying that group differences in the latent depression mean are meaningful despite the use of different instruments [59]. In practical terms, this means we can compare average depression levels between countries without confounding by measurement artefacts.

These findings extend prior harmonisation work. For example, the CLOSER collaboration successfully applied factor-analytic harmonisation to psychological distress measures across six British cohorts (McElroy et al., 2020). Our results build on this by pushing beyond a single-country context to show that a more stringent, generalisable framework can work across diverse populations. Similarly, studies in other domains have highlighted the value of advanced statistical harmonisation for cross-national comparisons. For instance, [60] harmonised cognitive measures between the US and India, finding that linking items via CFA enabled valid cross-country comparisons of cognitive function. In the same spirit, [61] demonstrated that a depressive symptom scale (EURO-D) achieved (approximate) scalar invariance across 27 European countries, supporting its use for cross-cultural comparisons. Together, these examples underscore that harmonisation methods can facilitate joint analysis of cohorts in different nations, allowing researchers to disentangle true population differences from measurement bias.

A key step in our approach was the use of an AI-based semantic matching tool (Harmony) to aid item harmonisation. Prior work has shown that such NLP-based tools can greatly reduce the manual effort in harmonisation: they quickly calculate high-potential item matches, which experts can then verify [47]. In our case, Harmony helped identify candidate matches across English and Portuguese scales, streamlining the mapping process. Nevertheless, we emphasise that semantic similarity does not guarantee conceptual equivalence. For example, Harmony gave a moderate similarity score (cosine ≈ 0.52) to the pairs “not feeling worth much as a person” and “not feeling one’s life is worthwhile.” Although both involve the notion of worth, an expert might note that the former taps self-esteem or self-concept, whereas the latter reflects a sense of purpose or meaning [62]. Such nuances are not distinguished by the embedding algorithm. Therefore, we adopted a hybrid strategy: automated matching to flag likely pairs, followed by clinician review to adjudicate subtle conceptual differences. This balance allowed us to gain efficiency from the algorithm while preserving domain fidelity. It is worth noting that tools like Harmony depend on the quality of the input text and metadata: different translations, ambiguity in item wording, or context differences can affect semantic scores. In practice, researchers should interpret the AI suggestions in light of substantive knowledge, using them as a guide rather than a final determinant.

Our harmonisation framework opens the door to robust cross-national analyses of mental health trajectories. With a common latent metric for depression, we can investigate how symptom courses diverge between countries, and what contextual factors drive those differences. For example, the UK and Brazil adopted markedly different COVID-19 policies, and harmonised data will allow a direct comparison of how lockdown stringency or social support measures affected depression trends [63]. More broadly, harmonised measures can link mental health trajectories to country-specific variables such as economic indicators, healthcare access, or public health interventions. Beyond the COVID-19 context, the same approach could be applied to other collective stressors. For example, natural or manmade disasters are known to have long-term psychological impacts, often compounding one another [64,65]￼. By harmonising data from different nations affected by successive events (e.g. wildfires, floods, economic crises), researchers could compare resilience and vulnerability factors internationally. In sum, cross-cohort harmonisation provides a powerful tool to ask global questions about mental health determinants, policy effects, and recovery patterns.

We recognise several limitations with our approach. Although we achieved configural and metric invariance for anxiety, our cross-cohort scalar model for anxiety did not converge. This likely reflects the smaller number and greater heterogeneity of overlapping anxiety items available between ELSA-UK and ELSA-Brasil. Insufficient item coverage can preclude full invariance: with only a handful of anxiety items, the model lacked power to align thresholds across groups. This failure highlights a general point: successful harmonisation requires a reasonably large pool of well-matched items. When instruments are too dissimilar or sparse, one may only achieve partial invariance. In practice, partial invariance can sometimes be acceptable; with guidelines suggesting that having at least half of the indicators invariant may allow for meaningful comparisons [66]. In extreme cases, problematic items might need to be dropped or given less weight in scoring [67]. In our harmonisation, none of the depression items had to be omitted, but researchers should be prepared to iterate: dropping or adjusting items is a valid strategy when invariance is violated. Ultimately, our anxiety results illustrate the difficulty of retrospectively harmonising constructs that were measured inconsistently across studies.

Furthermore, it is important to acknowledge the assumptions and trade-offs in our approach. Our framework is anchored in a priori decisions about which items captured the same symptom domains in both countries. These judgments assumed that, despite different wordings and answer formats, each paired item tapped an equivalent underlying construct. We also collapsed response categories into binary indicators to allow direct pooling; this simplification undoubtedly sacrificed some information in exchange for comparability [68]. Thus, the validity of our harmonised scores depends critically on the soundness of these initial decisions. Future users of this framework should be transparent about such choices and consider sensitivity analyses (e.g., testing alternative coding schemes) when feasible.

Alternative harmonisation strategies may also be considered to address some of the limitations of our approach, though each comes with its own trade-offs and assumptions. For example, when item-level data are unavailable, integrative data analysis (IDA) can pool raw scores across studies by rescaling or linking composite measures [69]. Similarly, test equating and linking methods [70] rely on anchor items or external calibration samples to align scales, potentially reducing the reliance on expert-driven dichotomisation. Meta-analytic approaches [71] provide another option when only study-level results are available, though these limit the ability to model within-person change or test measurement invariance directly. Additionally, multiple-group item response theory (IRT) provides an alternative harmonisation approach by testing differential item functioning (DIF) to evaluate whether items function equivalently across groups, thereby placing respondents on a common latent scale [72,73]. While more granular at the item level than MGCFA, it requires strong unidimensionality assumptions and larger sample sizes, and thus comes with its own trade-offs.

In summary, our structured harmonisation framework proved efficient and robust for depression: despite different instruments and languages, we derived a common latent depression metric that behaves consistently. For our anxiety construct, we were able to detect that invariance was not met, thus avoiding forcibly harmonising conceptually inconsistent data. However, the challenge with converging anxiety underscores the limits of retrospective harmonisation, especially when instruments diverge. These issues would only multiply as more cohorts or countries enter the analysis, reducing the chance of finding matching items across all datasets.

Therefore, future studies would benefit from prospective harmonisation efforts: agreeing on core symptom domains and standardised item sets (e.g. using DSM-based criteria or well-validated global mental health modules) at the design phase. Adopting common measurement frameworks or shared modules across international studies would minimise data loss and enhance comparability from the outset. Until then, our results show that careful, methodical harmonisation, combining expert knowledge, NLP tools, and factor analysis, can nevertheless yield meaningful cross-national mental health metrics.

## Data Availability

All data produced in the present study are available via application to Dementia Platforms UK (DPUK).

https://github.com/cgmhc/cgmhc_harmonisation

## Acknowledgements

This study was funded by the National Institute of Mental Health – 1RF1MH134638. PFZ has received funds from Fundação de Amparo à Pesquisa do Estado de São Paulo (FAPESP, grant number 2024/17532-0). DF has received funds from Wellcome Leap 1kD. ELSA-Brasil was funded by National Council for Scientific and Technological Development (CNPq) (Wave 1: BA 01 06 021200; ES 01 06 0300-00; MG 01 06 0278-00; 01 06 0071-00; RS 01 06 0010-00; SP 01 06 0115-00; FAPESP 2020/01476-2). We would also like to acknowledge the CGMHC cohort PI’s for their involvement in the project.

