## Supplemental materials 1 for "A robust framework for harmonising health measures across international cohorts: Evidence from the COVID-19 pandemic"

**Supplementary Materials**

**Questionnaire: ELSA-Brasil**

| **Variable Name** | **Description** | **Values** |
| --- | --- | --- |
| phq_01_c1 | Little interest or little pleasure in doing things? | Never = 1; Several days = 2; More than half of the days = 3; Almost every day = 4 |
| phq_02_c1 | Have you felt down, depressed or without perspective? | Never = 1; Several days = 2; More than half of the days = 3; Almost every day = 4 |
| dass1_c1 | I found it hard to wind down (Achei difícil me acalmar) | Did not apply at all = 0; Applied to some degree, or for a short time = 1; Applied to a considerable degree, or for a large part of the time = 2; Applied a lot, or most of the time = 3 |
| dass2_c1 | I was aware of dryness of my mouth (Senti minha boca seca) | Did not apply at all = 0; Applied to some degree, or for a short time = 1; Applied to a considerable degree, or for a large part of the time = 2; Applied a lot, or most of the time = 3 |
| dass3_c1 | I couldn't seem to experience any positive feeling at all (Não consegui vivenciar nenhum sentimento positivo) | Did not apply at all = 0; Applied to some degree, or for a short time = 1; Applied to a considerable degree, or for a large part of the time = 2; Applied a lot, or most of the time = 3 |
| dass4_c1 | I experienced breathing difficulty (eg, excessively rapid breathing,  breathlessness in the absence of physical exertion) Tive dificuldade em respirar em alguns momentos (ex. respiração ofegante, falta de ar, sem ter feito nenhum esforço físico)) | Did not apply at all = 0; Applied to some degree, or for a short time = 1; Applied to a considerable degree, or for a large part of the time = 2; Applied a lot, or most of the time = 3 |
| dass5_c1 | I found it difficult to work up the initiative to do things (Achei difícil ter iniciativa para fazer as coisas) | Did not apply at all = 0; Applied to some degree, or for a short time = 1; Applied to a considerable degree, or for a large part of the time = 2; Applied a lot, or most of the time = 3 |
| dass6_c1 | I tended to over-react to situations (Tive a tendência de reagir de forma exagerada às situações) | Did not apply at all = 0; Applied to some degree, or for a short time = 1; Applied to a considerable degree, or for a large part of the time = 2; Applied a lot, or most of the time = 3 |
| dass7_c1 | I experienced trembling (eg, in the hands) (Senti tremores (ex. nas mãos)) | Did not apply at all = 0; Applied to some degree, or for a short time = 1; Applied to a considerable degree, or for a large part of the time = 2; Applied a lot, or most of the time = 3 |
| dass8_c1 | I felt that I was using a lot of nervous energy (Senti que estava sempre nervoso) | Did not apply at all = 0; Applied to some degree, or for a short time = 1; Applied to a considerable degree, or for a large part of the time = 2; Applied a lot, or most of the time = 3 |
| dass9_c1 | I was worried about situations in which I might panic and make  a fool of myself (Preocupei-me com situações em que eu pudesse entrar em pânico e parecesse ridículo) | Did not apply at all = 0; Applied to some degree, or for a short time = 1; Applied to a considerable degree, or for a large part of the time = 2; Applied a lot, or most of the time = 3 |
| dass10_c1 | I felt that I had nothing to look forward to (Senti que não tinha nada a desejar) | Did not apply at all = 0; Applied to some degree, or for a short time = 1; Applied to a considerable degree, or for a large part of the time = 2; Applied a lot, or most of the time = 3 |
| dass11_c1 | I found myself getting agitated (Senti-me agitado) | Did not apply at all = 0; Applied to some degree, or for a short time = 1; Applied to a considerable degree, or for a large part of the time = 2; Applied a lot, or most of the time = 3 |
| dass12_c1 | I found it difficult to relax (Achei difícil relaxar) | Did not apply at all = 0; Applied to some degree, or for a short time = 1; Applied to a considerable degree, or for a large part of the time = 2; Applied a lot, or most of the time = 3 |
| dass13_c1 | I felt down-hearted and blue (Senti-me depressivo (a) e sem ânimo) | Did not apply at all = 0; Applied to some degree, or for a short time = 1; Applied to a considerable degree, or for a large part of the time = 2; Applied a lot, or most of the time = 3 |
| dass14_c1 | I was intolerant of anything that kept me from getting on with  what I was doing (Fui intolerante com as coisas que me impediam de continuar o que eu estava fazendo) | Did not apply at all = 0; Applied to some degree, or for a short time = 1; Applied to a considerable degree, or for a large part of the time = 2; Applied a lot, or most of the time = 3 |
| dass15_c1 | I felt I was close to panic (Senti que ia entrar em pânico) | Did not apply at all = 0; Applied to some degree, or for a short time = 1; Applied to a considerable degree, or for a large part of the time = 2; Applied a lot, or most of the time = 3 |
| dass16_c1 | I was unable to become enthusiastic about anything (Não consegui me entusiasmar com nada) | Did not apply at all = 0; Applied to some degree, or for a short time = 1; Applied to a considerable degree, or for a large part of the time = 2; Applied a lot, or most of the time = 3 |
| dass17_c1 | I felt I wasn't worth much as a person (Senti que não tinha valor como pessoa) | Did not apply at all = 0; Applied to some degree, or for a short time = 1; Applied to a considerable degree, or for a large part of the time = 2; Applied a lot, or most of the time = 3 |
| dass18_c1 | I felt that I was rather touchy (Senti que estava um pouco emotivo/sensível demais) | Did not apply at all = 0; Applied to some degree, or for a short time = 1; Applied to a considerable degree, or for a large part of the time = 2; Applied a lot, or most of the time = 3 |
| dass19_c1 | I was aware of the action of my heart in the absence of physical  exertion (eg, sense of heart rate increase, heart missing a beat) (Senti que meu coração estava alterado mesmo não tendo feito nenhum esforço físico) | Did not apply at all = 0; Applied to some degree, or for a short time = 1; Applied to a considerable degree, or for a large part of the time = 2; Applied a lot, or most of the time = 3 |
| dass20_c1 | I felt scared without any good reason (Senti medo sem motivo) | Did not apply at all = 0; Applied to some degree, or for a short time = 1; Applied to a considerable degree, or for a large part of the time = 2; Applied a lot, or most of the time = 3 |
| dass21_c1 | I felt that life was meaningless (Senti que a vida não tinha sentido) | Did not apply at all = 0; Applied to some degree, or for a short time = 1; Applied to a considerable degree, or for a large part of the time = 2; Applied a lot, or most of the time = 3 |

**Questionnaire: ELSA-UK**

| cvhospcncl_final010 | Hospital operation or treatment cancelled since outbreak: Mental health including depression (coded) | Yes / No |
| --- | --- | --- |
| CvMhCed_CvMhCed1_q | Much of the time during the past week: You felt depressed | Yes / No |
| CvMhCed_CvMhCed2_q | Much of the time during the past week: You felt that everything you did was an effort | Yes / No |
| CvMhCed_CvMhCed3_q | Much of the time during the past week: Your sleep was restless | Yes / No |
| CvMhCed_CvMhCed4_q | Much of the time during the past week: You were happy | Yes / No |
| CvMhCed_CvMhCed5_q | Much of the time during the past week: You felt lonely | Yes / No |
| CvMhCed_CvMhCed6_q | Much of the time during the past week: You enjoyed life | Yes / No |
| CvMhCed_CvMhCed7_q | Much of the time during the past week: You could not get going | Yes / No |
| CvMhCed_CvMhCed8_q | Much of the time during the past week: you felt sad | Yes / No |
| CvMhcaspGridB_CvMhcasp7_q | How often do you feel like: I look forward to each day | 1-4 |
| CvMhcaspGridB_CvMhcasp8_q | How often do you feel like: I feel that my life has meaning | 1-4 |
| CvMhcaspGridB_CvMhcasp9_q | How often do you feel like: I enjoy the things that I do | 1-4 |
| CvMhcaspGridB_CvMhcasp10_q | How often do you feel like: I feel full of energy these days | 1-4 |
| CvMhcaspGridB_CvMhcasp11_q | How often do you feel like: I feel that life is full of opportunities | 1-4 |
| CvMhcaspGridB_CvMhcasp12_q | How often do you feel like: I feel that the future looks good for me | 1-4 |
| CvMhgad_CvMhgad1_q | Over the last 2 weeks: Feeling nervous, anxious or on edge | 0-3 |
| CvMhgad_CvMhgad2_q | Over the last 2 weeks: Not being able to stop or control worrying | 0-3 |
| CvMhgad_CvMhgad3_q | Over the last 2 weeks: Worrying too much about different things | 0-3 |
| CvMhgad_CvMhgad4_q | Over the last 2 weeks: Trouble relaxing | 0-3 |
| CvMhgad_CvMhgad5_q | Over the last 2 weeks: Being so restless that it is hard to sit still | 0-3 |
| CvMhgad_CvMhgad6_q | Over the last 2 weeks: Becoming easily annoyed or irritable | 0-3 |
| CvMhgad_CvMhgad7_q | Over the last 2 weeks: Feeling afraid as if something awful might happen | 0-3 |
| CvMhOnsA | On a scale of 0 to 10, where 0 is "not at all" and 10 is "very", how happy, overall, did you feel yesterday? | 0-10 |
| CvMhOnsB | On a scale of 0 to 10, where 0 is "not at all" and 10 is "very", how anxious, overall, did you feel yesterday? | 0-10 |
| CvMhOnsC | On a scale of 0 to 10, where 0 is "not at all" and 10 is "very", how satisfied are you with your life nowadays? | 0-10 |
| CvMhOnsD | On a scale of 0 to 10, where 0 is "not at all" and 10 is "very", to what extent do you feel the things you do in your life are worthwhile? | 0-10 |
